# Development of machine learning models for the prediction of complications after colorectal and small intestine surgery in psychiatric and non-psychiatric patient collectives (P-Study)

**DOI:** 10.1101/2022.02.02.22269622

**Authors:** Stephanie Taha-Mehlitz, Bassey Enodien, Vincent Ochs, Ahmad Hendie, Anas Taha

## Abstract

**Introduction:** Psychiatric and psychosomatic diseases are an increasingly cumbersome burden for the medical system. Indeed, hospital costs associated with mental health conditions have been constantly on the rise in recent years. Moreover, psychiatric conditions are likely to have a negative effect on the treatment of other medical conditions and surgical outcomes, in addition to their direct effects on the overall quality of life. Our study aims to investigate the impact of preoperative risk factors, psychiatric and psychosomatic diseases, and non-psychiatric and non-psychosomatic diseases on the outcomes of small and large bowel surgery and length of hospital stay via predictive modeling techniques.

**Methods and Analysis:** Patient data will be collected from several participating national and international surgical centers. The machine learning models will be calculated and coded, but also published in respect to the TRIPOD guidelines (transparent reporting of a multivariable prediction model for individual prognosis or diagnosis).

**Expected Results:** It is conceivable to arrive at generalizable models predicting the above-mentioned endpoints through large amounts of data from several centers. The models will be subsequently deployed as a free-to-use web-based prediction tool.

**Ethics and Dissemination:** The ethical is approved by Cantonal Ethics Committee Zurich, Switzerland BASEC Nr. 2021-02105.

## 1. Background

Psychiatric and psychosomatic diseases are an increasingly cumbersome burden for the medical system. Indeed, hospital costs associated with mental health conditions have been constantly on the rise in recent years. Moreover, psychiatric conditions are likely to have a negative effect on the treatment of other medical conditions and surgical outcomes, in addition to their direct effects on the overall quality of life [1, 2].

Colorectal anastomotic insufficiencies (AI) and other complications after surgery cause a substantial clinical and economic burden on patients and can lead to a significant increase in the morbidity and mortality rate [3]. Hospital stays are significantly prolonged for patients who experience AI [3]. In expert centers, the estimated incidence of AI after colon anastomosis is at around 3.3%, and 8.6% for colorectal anastomoses [4]. In the literature, several somatic risk factors for the occurrence of anastomotic insufficiency have been described [5-8]. In contrast to this, and to the best of our knowledge, no studies are available that evaluate the influence of psychological risk factors on postoperative complications and anastomotic insufficiency after colorectal and small intestine surgery so far [9].

Our study aims to lay the basis for a predictive modeling service for risk factors, postoperative complications and prolonged hospital stay in patients suffering from psychiatric/psychosomatic and non-psychiatric/psychosomatic diseases undergoing small bowel and colorectal surgery.

## 1. Method

Patient data will be collected from several participating national and international surgical participants (hospitals, centers, university centers) in a database. The models will be built and published according to the TRIPOD guidelines [10].

### Ethical Approval

Every participating hospital is responsible for their own IRB approval (ethics board/institutional review board). After acquiring approval, the centers should collect retrospective and/or prospective data and share that data with the sponsor in a completely deidentified manner. The study protocol can be used as an assistance, which the sponsor will register on ClinicalTrials.gov.

Dr. med Anas Taha is the sponsor of the study. Dr. med. Bassey Enodien and Dr. Stephanie Taha-Mehlitz are the principal investigators.

### Authorship

Data of 500 patients (or more) which are complete, must be provided by each participating center. Each cooperating center is allowed to assign two center leads, and up to further two center-specific contributors will be stated as a contributor in PubMed/ Medline in a collaborative authorship model.

### Inclusion and Exclusion Criteria

Patients who underwent surgery with a colon, colorectal or small intestine anastomosis for several indications will be included like ileus, inflammatory bowel disease, diverticular disease, neoplasia or ischemia. Patients which are underaged, suffering from bear peritoneal carcinomatosis, recurrent colorectal cancer bear peritoneal carcinomatosis or unresectable metastatic disease at the time of surgery and anastomosis will be excluded.

Patients who are unable to provide informed approval to participate according to each center’s rules will be excluded. Furthermore, if patients cannot be followed up on for more than three months after surgery, they will be excluded because the occurrence of postoperative complications can neither be confirmed nor excluded.

### Data Collection

The data will be assembled and processed by each center from their dedicated clinical database before submitting them to the study-specific REDCap database provided by the sponsor. Each participating center must provide data of a minimum of 500 patients. Patient data will be collected between January 2012 and December 2021. To ensure anonymity, the entries will be stored in a completely deidentified manner. Upon completion of data entrance of each patient, a study-specific patient identifier will be produced. The final data on the database will not contain any identifiable patient information. To ensure that processed patient data can be re-evaluated, each center should maintain a log file for each specific patient in the study.

### Primary Endpoint Definition

The following three study endpoints will be assessed separately.

1. Anastomotic insufficiency/leakage as defined by Gessler et al. [8] and Rahbari et al. [11]. Predictive model with an app for the prediction of AI based on risk factors.
2. Comprehensive Complication Index as defined by Slankamenac et al., 2013 [12].
3. Length of Hospital Stay, defined as time span in days from surgery to discharge from the service. And whether the patients who suffer from psychiatric and psychosomatic disorders are having higher complication rates and longer length of hospital stay.
4. How does the intraoperative fluid management play a role on the development of an AI?
5. Development of a preoperative score to better assess the patient’s condition and find patients at risk for morbidity/mortality in small bowel and colorectal surgery.

### Features

- Age [13]
- Gender [14]
- Body mass index [15]
- Height
- Weight [16]
- Charlson Comorbidity Index (CCI) [17]
- Nutritional status [18]
- American Society of Anesthesiologists Score (ASA) [19]
- Smoking status [20]
- Alcohol abuse status [21]
- Kidney function [22]
- Surgical indication [23]
- Type of operation [24]
- Previous abdominal surgery
- Emergency [25]
- Approach to the abdominal cavity [26]
- Anastomotic technique [27]
- Defunctioning stoma [28]
- TNM classification [29]
- Hepatic metastasis [30]
- Distance from anal verge [31]
- Preoperative leucocyte count [32]
- Preoperative albumin count [33]
- Preoperative hemoglobin level [34]
- Preoperative steroid use [35]
- Preoperative radiotherapy and/or chemotherapy [36]
- Psychiatric and psychosomatic diseases according to the ICD-10 classification
- Depression
- Schizophrenia
- Bipolar disorder
- Obsessive-compulsive disorder
- Anxiety and panic disorder
- Anorexia, bulimia, and other eating disorders
- Dementia
- History of drug abuse (harmful use, drug addiction, withdrawal syndrome, anda ssociated long-term consequences.

### Assessment for postoperative complications

The complication rate will be evaluated after colorectal surgery, and the Comprehensive Complication Index (CCI) [12, 37] will be calculated for each patient. Furthermore, the occurrence of AI (as defined by Gessler [8] and Rahbari [11]), length of hospital stay (LOS), mortality rate and readmission rate will be assessed and correlated to the occurrence of psychiatric and psychosomatic diseases.

### Sample Size

There are two main problems associated with predictive modeling. The first of these is the relevance of the features used for model creation. Indeed, it is impossible to detect a relationship between a set of features and the outcome variable when there is none. Logically, this problem cannot be overcome even with the most sophisticated algorithms. Therefore, a comprehensive literature study was necessary to ensure that the risk factors listed as features above have been consistently reported as risk factors for postoperative complications after small bowel and colorectal surgery. Admittedly, very little literature is available connecting psychiatric and psychosomatic, but also non-psychiatric and non-psychosomatic diseases to adverse postoperative outcomes. Our group is currently aiming to fill this gap with a retrospective study that aims to clarify the putative connection between mental health and postoperative complications. The second important point is the sample size, since the relationship between the prediction performance and the sample size is almost directly proportional. There are three important considerations to be made when determining a suitable sample size. First, the sample size should be large enough to ensure that the sample is representative enough for the study population of interest. Second, the sample size should be appropriate for the algorithm used. For instance, deep neural networks require thousands of data sets to assemble a model, while stable results can be accomplished by a logistic regression model using only a few hundred data sets. Third, the number of input variables is proportional to the number of data sets necessary for a useful prediction. As an established rule of thumb, at least teen or more positive cases are required per input variable which have been included to model the relationships. Consequently, very rare outcomes need much larger data sets than common outcomes. For example, given a 10% occurrence rate for the outcome, a prediction model with fifteen or more input variables would likely require a minimum of 150 positive cases and 1500 cases total. Furthermore, larger sample sizes usually allow for enhanced training and validation sets and thus more reliable and generalizable results.

In the current study, a total of at least 13 different features were selected. Consequently, at least 130 patients with the desirable outcome will be necessary. However, to ensure the quality of our study through extensive model calibration, a goal of 300 patients accounting for the outcome variable was set. With a frequency of specific postoperative complications like AI of about 2% in small intestine anastomosis, 3% in colon- and 9% in colorectal anastomosis, a total of 7,500 as a target value for the average incidence of complications will be necessary to create a high-performing model. To allow for adequate evaluation of calibration and validation we estimate further 2,500 patient datasets for a test set. Consequently, a total number of 10,000 patient data sets will be necessary for this study. However, greater performance and better calibration will be achieved by applying more data.

### Predictive Modeling

Missing data will be tolerated up to a margin of 25% per patient or feature. A concomitantly trained nearest neighbor imputer will be used to impute missing data up to the above-mentioned margin [38]. Features and patients lacking more than 25% of the data will be systematically excluded. If major outcome imbalances are discovered [39, 40], upsampling will be performed on the training set using either random upsampling or synthetic minority oversampling (SMOTE). Furthermore, if necessary, recursive variables elimination (RFE) will be employed for feature selection on the training set [41].

The following algorithms will be trialed for binary classification: the stochastic gradient boosting machine (GBM), random forest, generalized additive model (GAM), artificial neural network, the generalized linear model (GLM), support vector machine (SVM), and naïve Bayes classifier. Each model will be fully trained and hyperparameter tuned where applicable. For the best-performing model, the resampled training performance will be examined.

The model with the best performance in the training set will subsequently be examined on the test data set for external validation. A bootstrap of the test data will be used to calculate the 95% confidence intervals (CI) for the external validation metrics.

The binary classification threshold will either be established on the training data alone using the AUC-based “closest-to-(0,1)-criterion” or Youden’s index. For the analyses, Python Version 3.7.12 will be used [42 - 44].

### Evaluation

Model discrimination and calibration are the most common metrics applied to evaluate classification models [45]. Discrimination describes the procedure of a model to correctly identify and assign binary problems. This means its ability to correctly predict whether or not a certain outcome will occur. In comparison, calibration describes how precisely the continuous probabilities (probability range from zero percentage to one-hundred) from a model correspond to the observed true occurrence of a binary outcome. Even though calibration metrics are scarcely reported in publications, they are much appreciated by clinicians and patients as they allow for a more figurative description of the patient’s risk [45].

For calibration as well as for discrimination, the resampled training performance and the external validation performance will be evaluated. For discrimination performance, we will use the following metrics: AUC, accuracy, recall, precision, positive predictive value (PPV), negative predictive value (NPV), and F1 Score will be assessed. For calibration metrics, the following metrics will be used: the Brier score, expected-observed (E/O) ratio and the calibration slope and intercept.

### Interpretability

The degree of interpretability of this study’s results will be based on the best-performing algorithm. While certain algorithms by design provide easily interpretable insights into the effect that features have on the outcome (e.g., Random forest), more complex models such as neural networks or stochastic gradient boosting machines cannot provide definite explanations for their results. For the latter group of algorithms, an AUC-based variable importance and the LIME principle will be used to provide a model-agnostic local interpretation of variable importance [46].

### 2. Expected Results

It is conceivable to arrive at generalizable models predicting the above-mentioned endpoints through large amounts of data from several centers. The models will be subsequently deployed as a free-to-use web-based prediction tool. The sponsor will cover the cost of the hosting server and digital infrastructure. The sponsor will store the data for ten years. The model and the data will be stored by the sponsor for ten years.

## Data Availability

All data produced in the present work are contained in the manuscript

## Author Contributions

Conceptualization A.T.; S. T.M. administration and ethics. A. T.; B. E.; S.T.-M.; writing—original draft preparation A.T., S.T.-M.; writing—review and editing S.T.-M., V.O., A.H.; All authors have read and agreed to the published version of the manuscript.

## Acknowledgment

Marta Bachman helped with the ethics.

## Literature

1. Mavros, M.N., et al., Do psychological variables affect early surgical recovery? PLoS One, 2011. 6(5): p. e20306.

2. Barrett-Bernstein, M., et al., Depression and functional status in colorectal cancer patients awaiting surgery: Impact of a multimodal prehabilitation program. Health Psychol, 2019. 38(10): p. 900–909.

3. Lee, S.W., D. Gregory, and C.L. Cool, Clinical and economic burden of colorectal and bariatrica nastomotic leaks. Surg Endosc, 2020. 34(10): p. 4374–4381.

4. Krell, R.W., et al., Hospital readmissions after colectomy: a population-based study. J Am CollSurg, 2013. 217(6): p. 1070–9.

5. Konishi, T., et al., Risk factors for anastomotic leakage after surgery for colorectal cancer: results of prospective surveillance. J Am Coll Surg, 2006. 202(3): p. 439–44.

6. Slieker, J.C., et al., Long-term and perioperative corticosteroids in anastomotic leakage: a prospective study of 259 left-sided colorectal anastomoses. Arch Surg, 2012. 147(5): p. 447–52.

7. Silva-Velazco, J., et al., Is there anything we can modify among factors associated with morbidity following elective laparoscopic sigmoidectomy for diverticulitis? Surg Endosc, 2016.30(8): p. 3541–51.

8. Gessler, B., O. Eriksson, and E. Angenete, Diagnosis, treatment, and consequences of anastomotic leakage in colorectal surgery. Int J Colorectal Dis, 2017. 32(4): p. 549–556.

9. Hijazi, Y., U. Gondal, and O. Aziz, A systematic review of prehabilitation programs ina bdominal cancer surgery. Int J Surg, 2017. 39: p. 156–162.

10. Collins, G.S., et al., Protocol for development of a reporting guideline (TRIPOD-AI) and risk of bias tool (PROBAST-AI) for diagnostic and prognostic prediction model studies based on artificial intelligence. BMJ Open, 2021. 11(7): p. e048008.

11. Rahbari, N.N., et al., Definition and grading of anastomotic leakage following anterior resection of the rectum: a proposal by the International Study Group of Rectal Cancer.Surgery, 2010. 147(3): p. 339–51.

12. Slankamenac, K., et al., The comprehensive complication index: a novel continuous scale to m easure surgical morbidity. Ann Surg, 2013. 258(1): p. 1–7.

13. Kumar, Ashok. “Anterior Resection for Rectal Carcinoma -Risk Factors for Anastomotic Leaksand Strictures.” World Journal of Gastroenterology, vol. 17, no. 11, 2011, p. 1475, 10.3748/wjg.v17.i11.1475.

14. Trencheva, K., et al., Identifying important predictors for anastomotic leak after colon and rectal resection: prospective study on 616 patients. Ann Surg, 2013. 257(1): p. 108–13.

15. Komen, Niels, et al. “After-Hours Colorectal Surgery: A Risk Factor for Anastomotic Leakage.” International Journal of Colorectal Disease, vol. 24, no. 7, 21 Mar. 2009, pp. 789– 795, 10.1007/s00384-009-0692-4.

16. Rullier, E, et al. “Risk Factors for Anastomotic Leakage after Resection of Rectal Cancer.” British Journal of Surgery, vol. 85, no. 3, Mar. 1998, pp. 355–358, 10.1046/j.1365-2168.1998.00615.x.

17. Tian, Yaohua, et al. “Comorbidity and the Risk of Anastomotic Leak in Chinese Patients with Colorectal Cancer Undergoing Colorectal Surgery.” International Journal of Colorectal Disease, vol. 32, no. 7, 23 Mar. 2017, pp. 947–953, 10.1007/s00384-017-2798-4.

18. Lee, Soo Young, et al. “Nutritional Risk Screening Score Is an Independent Predictive Factor of Anastomotic Leakage after Rectal Cancer Surgery.” European Journal of Clinical Nutrition, vol. 72, no. 4, 19 Feb. 2018, pp. 489–495, 10.1038/s41430-018-0112-3.

19. Kryzauskas, Marius, et al. “Risk Factors for Anastomotic Leakage and Its Impact on Long-Term Survival in Left-Sided Colorectal Cancer Surgery.” World Journal of Surgical Oncology, vol. 18, 1 4 Aug. 2020, www.ncbi.nlm.nih.gov/pmc/articles/PMC7427291/,10.1186/s12957-020-01968-8.

20. Fawcett, A, et al. “Smoking, Hypertension, and Colonic Anastomotic Healing; a Combined Clinical and Histopathological Study.” Gut, vol. 38, no. 5, 1 May 1996, pp. 714–718, 10.1136/gut.38.5.714.

21. Sørensen, L. T., et al. “Smoking and Alcohol Abuse Are Major Risk Factors for Anastomotic Leakage in Colorectal Surgery.” British Journal of Surgery, vol. 86, no. 7, 1 July 1999, pp. 927–9 31, 10.1046/j.1365-2168.1999.01165.x.

22. Wako, Gutema, et al. “Colorectal Anastomosis Leak: Rate, Risk Factors and Outcome in a Tertiary Teaching Hospital, Addis Ababa Ethiopia, a Five Year Retrospective Study.” Ethiopian Journal of Health Sciences, vol. 29, no. 6, 1 Nov. 2019, 10.4314/ejhs.v29i6.14.

23. Platell, C., et al. “The Incidence of Anastomotic Leaks in Patients Undergoing Colorectal Surgery.” Colorectal Disease: The Official Journal of the Association of Coloproctology of Great Britain and Ireland, vol. 9, no. 1, 1 Jan. 2007, pp. 71–79, pubmed.ncbi.nlm.nih.gov/17181849/, 10.1111/j.1463-1318.2006.01002.x.

24. Law, Wai Lun, and Kin Wah Chu. “Anterior Resection for Rectal Cancer with Mesorectal Excision.” Annals of Surgery, vol. 240, no. 2, Aug. 2004, pp. 260–268, 10.1097/01.sla.0000133185.23514.32.

25. Awad, Selmy, et al. “The Assessment of Perioperative Risk Factors of Anastomotic Leakageafter Intestinal Surgeries; a Prospective Study.” BMC Surgery, vol. 21, no. 1, 7 Jan. 2021, 10.1186/s12893-020-01044-8.

26. Vennix, Sandra, et al. “Laparoscopic versus Open Total Mesorectal Excision for Rectal Cancer.” Cochrane Database of Systematic Reviews, 15 Apr. 2014, 10.1002/14651858.cd005200.pub3.

27. Boccola, Mark A., et al. “Risk Factors and Outcomes for Anastomotic Leakage in Colorectal Surgery: A Single-Institution Analysis of 1576 Patients.” World Journal of Surgery, vol. 35, no.1, 23 Oct. 2010, pp. 186–195, 10.1007/s00268-010-0831-7.

28. Jestin, P., et al. “Risk Factors for Anastomotic Leakage after Rectal Cancer Surgery: A Case-Control Study.” Colorectal Disease, vol. 10, no. 7, 21 Aug. 2008, pp. 715–721, 10.1111/j.1463-1318.2007.01466.x.

29. Yeh, Chien Yuh, et al. “Pelvic Drainage and Other Risk Factors for Leakage after Elective Anterior Resection in Rectal Cancer Patients.” Annals of Surgery, vol. 241, no. 1, Jan. 2005, pp.9 –13, 10.1097/01.sla.0000150067.99651.6a.

30. Sakr, Ahmad, et al. “Predictive Factors for Small Intestinal and Colonic Anastomotic Leak: A Multivariate Analysis.” Indian Journal of Surgery, vol. 79, no. 6, 17 Oct. 2016, pp. 555–562, 10.1007/s12262-016-1556-0.

31. Matthiessen, P., et al. “Risk Factors for Anastomotic Leakage after Anterior Resection of the Rectum.” Colorectal Disease, vol. 6, no. 6, Nov. 2004, pp. 462–469, 10.1111/j.1463-1318.2004.00657.x.

32. Morimoto, Masaki, et al. “Preoperative White Blood Cell Count Predicts Anastomotic Leakage in Patients with Left-Sided Colorectal Cancer.” PLOS ONE, vol. 16, no. 10, 20 Oct. 2021, p. e0258713, 10.1371/journal.pone.0258713.

33. Telem, Dana A. “Risk Factors for Anastomotic Leak Following Colorectal Surgery.” Archives of Surgery, vol. 145, no. 4, 1 Apr. 2010, p. 371, 10.1001/archsurg.2010.40.

34. Yeap, Evie, et al. “Preoperative Anaemia and Thrombocytopenia Are Associated with Venous Thromboembolism Complications after Colorectal Resection.” ANZ Journal of Surgery, vol. 91,no. 1-2, May 2020, 10.1111/ans.15918.

35. Golub, R., et al. “A Multivariate Analysis of Factors Contributing to Leakage of Intestinal Anastomoses.” Journal of the American College of Surgeons, vol. 184, no. 4, 1 Apr. 1997, pp.3 64–372, pubmed.ncbi.nlm.nih.gov/9100681/.

36. Martel, Guillaume et al. “Neoadjuvant therapy and anastomotic leak after tumor-specific mesorectal excision for rectal cancer.” Diseases of the colon and rectum vol. 51,8 (2008):1195–201. doi:10.1007/s10350-008-9368-3

37. Slankamenac, K., et al., The comprehensive complication index: a novel and more sensitive endpoint for assessing outcome and reducing sample size in randomized controlled trials. AnnSurg, 2014. 260(5): p. 757–62; discussion 762-3.

38. Templ, M., et al., VIM: Visualization and Imputation of Missing Values, 2012. 2019. 1(0).

39. Chawla, N.V., et al., SMOTE: synthetic minority over-sampling technique. 2002. 16: p. 321–357.

40. Staartjes, V.E. and M.L.J.J.o.N.S. Schröder, Letter to the Editor. Class imbalance in machine learning for neurosurgical outcome prediction: are our models valid? 2018. 29(5): p. 611–612.

41. Granitto, P.M., et al., Recursive feature elimination with random forest for PTR-MS analysis ofagroindustrial products. 2006. 83(2): p. 83–90.

42. Bakker, I.S., et al., Risk factors for anastomotic leakage and leak-related mortality aftercolonic cancer surgery in a nationwide audit. Br J Surg, 2014. 101(4): p. 424–32.

43. Buchs, N.C., et al., Incidence, consequences, and risk factors for anastomotic dehiscence afterc olorectal surgery: a prospective monocentric study. Int J Colorectal Dis, 2008. 23(3): p. 265–70.

44. Choi, H.K., W.L. Law, and J.W. Ho, Leakage after resection and intraperitoneal anastomosis for colorectal malignancy: analysis of risk factors. Dis Colon Rectum, 2006. 49(11): p. 1719–25.

45. Staartjes, V.E. and J.M.J.J.o.N.S. Kernbach, Letter to the Editor. Importance of calibrationassessment in machine learning–based predictive analytics. 2020. 32(6): p. 985–987.

46. Ribeiro, M.T., S. Singh, and C. Guestrin. “ Why should i trust you?” Explaining the predictionso f any classifier. in Proceedings of the 22nd ACM SIGKDD international conference on knowledge discovery. and data mining. 2016

